# Vitamin D and COVID-19 susceptibility and severity in the COVID-19 Host Genetics Initiative: A Mendelian randomization study

**DOI:** 10.1101/2020.09.08.20190975

**Authors:** Guillaume Butler-Laporte, Tomoko Nakanishi, Vincent Mooser, David R. Morrison, Tala Abdullah, Olumide Adeleye, Noor Mamlouk, Nofar Kimchi, Zaman Afrasiabi, Nardin Rezk, Annarita Giliberti, Alessandra Renieri, Yiheng Chen, Sirui Zhou, Vincenzo Forgetta, J Brent Richards

## Abstract

**Background:** Increased vitamin D levels, as reflected by 25OHD measurements, have been proposed to protect against COVID-19 disease based on *in-vitro*, observational, and ecological studies. However, vitamin D levels are associated with many confounding variables and thus associations described to date may not be causal. Vitamin D Mendelian randomization (MR) studies have provided results that are concordant with large-scale vitamin D randomized trials. Here, we used two-sample MR to assess evidence supporting a causal effect of circulating 25OHD levels on COVID-19 susceptibility and severity.

**Methods and findings:** Genetic variants strongly associated with 25OHD levels in a genome-wide association study (GWAS) of 443,734 participants of European ancestry (including 401,460 from the UK Biobank) were used as instrumental variables. GWASs of COVID-19 susceptibility, hospitalization, and severe disease from the COVID-19 Host Genetics Initiative were used as outcome GWASs. These included up to 14,134 individuals with COVID-19, and 1,284,876 without COVID-19, from 11 countries. SARS-CoV-2 positivity was determined by laboratory testing or medical chart review. Population controls without COVID-19 were also included in the control groups for all outcomes, including hospitalization and severe disease. Analyses were restricted to individuals of European descent when possible. Using inverse-weighted MR, genetically increased 25OHD levels by one standard deviation on the logarithmic scale had no significant association with COVID-19 susceptibility (OR = 0.97; 95% CI: 0.95, 1.10; P=0.61), hospitalization (OR = 1.11; 95% CI: 0.91, 1.35; P=0.30), and severe disease (OR = 0.93; 95% CI: 0.73, 1.17; P=0.53). We used an additional 6 meta-analytic methods, as well as sensitivity analyses after removal of variants at risk of horizontal pleiotropy and obtained similar results. These results may be limited by weak instrument bias in some analyses. Further, our results do not apply to individuals with vitamin D deficiency.

**Conclusion:** In this two-sample MR study, we did not observe evidence to support an association between 25OHD levels and COVID-19 susceptibility, severity, or hospitalization. Hence, vitamin D supplementation as a means of protecting against worsened COVID-19 outcomes is not supported by genetic evidence. Other therapeutic or preventative avenues should be given higher priority for COVID-19 randomized controlled trials.

**Author Summary:**

- **Why was this study done?**
  - Vitamin D levels have been associated with COVID-19 outcomes in multiple observational studies, though confounders are likely to bias these associations.
  - By using genetic instruments which limit such confounding, Mendelian randomization studies have consistently obtained results concordant with vitamin D supplementation randomized trials. This provides rationale to undertake vitamin D Mendelian randomization studies for COVID-19 outcomes.
- **What did the researchers do and find?**
  - We used the genetic variants obtained from the largest consortium of COVID-19 cases and controls, and the largest study on genetic determinants of vitamin D levels.
  - We used Mendelian randomization to estimate the effect of increased vitamin D on COVID-19 outcomes, while limiting confounding.
  - In multiple analyses, our results consistently showed no evidence for an association between genetically predicted vitamin D levels and COVID-19 susceptibility, hospitalization, or severe disease.
- **What do these findings mean?**
  - Using Mendelian randomization to reduce confounding that has traditionally biased vitamin D observational studies, we did not find evidence that vitamin D supplementation in the general population would improve COVID-19 outcomes
  - These findings, together with recent randomized controlled trial data, suggest that other therapies should be prioritized for COVID-19 trials.

## Introduction

SARS-CoV-2 infection has killed millions of individuals and has led to the largest economic contraction since the Great Depression [1]. Therefore, therapies are required to treat severe COVID-19 disease and to prevent its complications. Therapeutic development, in turn, requires well-validated drug targets to lessen COVID-19 severity.

Recently, vitamin D status, as reflected by 25-hydroxy-vitamin D (25OHD) level has been identified as a potentially actionable drug target in the prevention and treatment of COVID-19 [2]. As the pre-hormone to the biologically active calcitriol, 25OHD has been epidemiologically linked to many health outcomes [3,4]. Given calcitriol’s recognized *in-vitro* immunomodulatory role [5], as well as observational and ecological studies associating measured 25OHD blood levels with COVID-19 [6,7], the vitamin D pathway might be a biologically plausible target in COVID-19. This could be of public health importance, given that the prevalence of vitamin D insufficiency is high in most countries, and that more than 37% of elderly adults in the USA take vitamin D supplements [8]. Further, 25OHD supplementation is inexpensive and reasonably safe—thus providing a potential avenue to lessen the burden of the SARS-CoV-2 pandemic.

However, observational studies on 25OHD are prone to confounding and reverse causation bias. Confounding happens when the relationship between exposure (25OHD) and the outcome (COVID-19) is influenced by unobserved, or improperly controlled common causes. Reverse causation happens when the outcome itself is a cause of the exposure. Likewise, conclusions drawn from *in-vitro* may not be applicable *in-vivo*. Accordingly, randomized controlled trials (RCTs) on 25OHD supplementation have been undertaken to test their effect on disease outcomes where observational studies have supported a role for 25OHD level. However, across endocrinology, respirology, cardiology, and other specialties, these trials have most often not demonstrated statistically significant benefits [9–11]. Some RCTs have even shown detriment to 25OHD supplementation [12]. In the field of infectious diseases, an individual patient data meta-analysis of randomized controlled trial of 25OHD supplementation [13] showed some benefit to prevent respiratory tract infections (OR 0.80, 95% CI: 0.69 to 0.93). However, this effect was driven by generally benign upper respiratory tract infections, was not observed in lower respiratory tract disease (OR: 0.96, 95% CI: 0.83 to 1.10) and even showed numerically worse all-cause mortality (OR: 1.39, 95% CI: 0.85 to 2.27). Likewise, a recent trial on sepsis obtained a numerically higher mortality rate in patients who received 25OHD supplementation [14]. At present, we are aware of two RCTs testing the role of vitamin D supplementation on COVID-19 outcomes, both using high-dose vitamin D given at time of hospital admission for COVID-19. The first [15] was a small trial (n=75) showing less intensive care unit admissions in the vitamin D treated arm. However, the follow-up time for mortality varied, and the open-label design put it at high risk of bias. The second [16] was a larger study (n=240) using a double-blind design, and showed no effect on mortality, risk of mechanical ventilation, and length of stay. Nevertheless, questions remain on the use of pre-illness vitamin D supplementation and its effect on disease susceptibility. While RCTs can control for confounding and provide unbiased estimates of the effect of 25OHD supplementation in COVID-19, large well-designed RCTs require considerable resources and time.

Mendelian randomization (MR) is a genetic epidemiology method that uses genetic variants as instrumental variables to infer the causal effect of an exposure (in this case 25OHD level) on an outcome (in this case, COVID-19 susceptibility and severity) [17]. MR overcomes confounding bias since genetic alleles are randomized to the individual at conception, thereby breaking associations with most confounders. Similarly, since genetic alleles are always assigned prior to disease onset, they are not influenced by reverse causation. MR has been used in conjunction with proteomics and metabolomics to prioritize drug development and repurposing, and support investment in RCTs which have a higher probability of success [18,19]. In the case of vitamin D, MR has been able to provide causal effect estimates consistently in line with those obtained from RCTs [9,20–24], or support the use of vitamin D supplementation in preventing diseases in at risk individuals (most notably multiple sclerosis [25]). Hence, MR may support investments in 25OHD supplementation trials in COVID-19, if a benefit was shown. Further, since MR results can be generated rapidly, such evidence may provide interim findings while awaiting RCT results.

However, MR relies on several core assumptions [26]. First, genetic variants must be associated with the exposure of interest. Second, they should not affect the outcome except through effects on the exposure (also known as lack of horizontal pleiotropy). Specifically, MR also assumes that the relationship between the exposure and the outcome is linear. However, this assumption still provides a valid test of the null hypothesis when studying population-level effects [27], as MR then measures the population-averaged effect on the outcome of a shift in the distribution of the exposure. Third, genetic variants should not associate with the confounders of the exposure-outcome relationship. Of these, the most problematic is the second assumption. Yet, in the case of 25OHD, many of its genetic determinants reside at loci that harbour genes whose roles in 25OHD production, metabolism and transport are well known [25]. Leveraging this known physiology can help to prevent the incorporation of genetic variants that could lead to horizontal pleiotropy.

Here, we used genetic determinants of serum 25OHD from a recent genome-wide association study (GWAS) and meta-analysis of more than 443,734 participants of European ancestry [28] in an MR study to test the relationship between increased 25OHD level and COVID-19 susceptibility and severity.

## Methods

We used a two-sample MR approach to estimate the effect of 25OHD levels on COVID-19 susceptibility and severity. In two-sample MR [29], the effect of genetic variants on 25OHD and on COVID-19 outcomes are estimated in separate GWASs from different populations. This allows for increased statistical power by increasing the sample size in both the exposure and outcome cohorts. This study is reported as per the Strengthening the Reporting of Observational Studies in Epidemiology (STROBE) guideline [30] (**Supplement 1**).

Our study did not employ a prospective protocol. Analyses were first planned and performed in July 2020 and updated following peer-review in December 2020. Three major changes were made during the update. First, we used the most up to date COVID-19 Host Genetics Initiative (COVID-19 HGI) GWAS summary statistics. These were made available during the peer-review process. Second, to alleviate potential selection and collider bias, we modified the outcome phenotypes to include population controls. We also performed additional MR sensitivity analyses to check for the robustness of our results. The latter two modifications were made at the request of peer-reviewers. Finally, minor changes to the results’ interpretations were made following further peer-review in February 2021.

### Choice of 25OHD genetic instruments

To find genetic variants explaining 25OHD levels [28], we used a GWAS from our group, which is the largest published GWAS of 25OHD levels, to the best of our knowledge. Importantly, this meta-analysis controlled for season of vitamin D measurement to obtain genetic variants significantly associated with 25OHD levels. From the list of conditionally independent variants provided, we further selected SNPs whose effect on 25OHD level was genome-wide significant (P<5×10^−8^), minor allele frequency was more than 1%, and with linkage disequilibrium coefficients (r^2^) of less than 5% (using the LDlink [31] tool and the European 1000 Genomes dataset, exclusing Finnish populations). For SNPs that were not available in the outcome GWAS or with palindromic alleles of intermediate frequency (between 42% and 58%), we again used the LDlink [31] tool to find genetic proxies in the European 1000 Genomes dataset (excluding Finnish populations) using linkage disequilibrium r^2^ of 90% or more.

### COVID-19 outcome definitions and GWASs

We used the COVID-19 HGI outcome definitions and GWAS summary statistics for COVID-19 susceptibility, hospitalization, and severe disease outcomes [32]. For all outcomes, a COVID-19 infection was defined as a positive SARS-CoV-2 laboratory test (e.g. RNA RT-PCR or serology tests) or electronic health record evidence of SARS-CoV-2 infection (using International Classification of Diseases or physician notes). The susceptibility phenotype compared COVID-19 cases, with controls which were defined as any individuals without a history of COVID-19. The hospitalized outcome compared cases defined as hospitalized patients with COVID-19, and controls as any individuals not experiencing a hospitalization for COVID-19, which includes those without COVID-19. The severe disease outcome cases were defined as hospitalized individuals with COVID-19 who required respiratory support. Respiratory support was defined as intubation, CPAP, BiPAP, continuous external negative pressure, or high flow nasal cannula. Controls for the severe COVID-19 outcome were defined as individuals without severe COVID-19 (including those without COVID-19). The inclusion of COVID-19 negative participants as controls in each outcome decreases the possibility of collider bias [33] and allows for better population level comparisons. These three outcome phenotypes are referred to as C2, B2, and A2, respectively, in the COVID-19 HGI documentation.

For our study, we used the October 20^th^ 2020 (v4) COVID-19 HGI fixed effect meta-analysis of GWAS from up to 22 cohorts, performed in up to 11 countries. Every participating cohort was asked to provide summary statistics from a GWAS on the above three outcomes, and including the following non-genetic covariates: age, sex, age*age, age*sex, 20 genetic principal components, as well as any locally relevant covariates at the discretion of participating studies (e.g. hospital, genotype panel, etc.). Cohorts were asked to follow common sample and variant quality control, and only performed analysis if they enrolled 100 cases or more. Analyses were done separately for each major ancestry group to further control for population stratification. For the purposes of our study, we used the meta-analysis results from European ancestry cohorts, except for the severe COVID-19 outcome, for which this meta-analysis was not available. Further details on the three phenotypes and participating cohorts are found in **Table 1** and **Supplement 2**.

**Table 1:** Sources of data for the analysis. COVID-19 susceptibility and severity outcomes are taken from the COVID-19 HGI. See **Supplement 2** for details on cohorts of COVID-19 susceptibility and severity phenotypes. 25OHD: 25-hydroxy vitamin D. GWAS: genome-wide association study. UKB: UK Biobank. CPAP: continuous positive airway pressure ventilation. BiPAP: bilevel positive airway pressure ventilation.

| Phenotype | Source of genetic variants |  |
| --- | --- | --- |
|  | Cohort | Participants |
| 25OHD circulating levels | Manousaki *et al* | Meta-analysis of two 25OHD GWAS:<br>- 401,460 adult white British participants from the UKB<br>- 42,274 from an international consortium of adult individuals of European ancestry |
| COVID-19 susceptibility | Susceptibility | Meta-analysis of 22 GWAS performed in individuals of European ancestry from 11 countries:<br>- **Cases:** 14,134 individuals with COVID-19 by laboratory confirmation, chart review, or self-report<br>- **Controls:** 1,284,876 individuals without confirmation or history of COVID-19 |
| COVID-19 severity | Hospitalized | Meta-analysis of 13 GWAS performed in individuals of European ancestry from 11 countries:<br>**Cases:** 6,406 hospitalized individuals with COVID-19<br>**Controls:** 902,088 without hospitalization with COVID-19 |
|  | Severe Disease | Meta-analysis of 12 GWAS performed in individuals of European ancestry from 9 countries:<br>- **Cases:** 4,336 SARS-CoV-2 infected hospitalized individuals who died or required respiratory support (intubation, CPAP, BiPAP, continuous external negative pressure, high flow nasal cannula).<br>- **Controls:** 623,902 without severe COVID-19 |

### Primary MR analysis

The effect of 25OHD level on COVID-19 outcomes was obtained for each SNP by using the Wald ratio method. The effect of each SNP was given in standardized log-transformed 25OHD level. Each estimate was meta-analyzed using the inverse-variance weighted (IVW) method, and we performed variant heterogeneity tests to check robustness of IVW results. Allele harmonization and computations were performed using the TwoSampleMR package [34].

### Horizontal pleiotropy sensitivity analysis

We undertook multiple analyses to assess the risk of horizontal pleiotropy (a violation of the second MR assumption). First, we used MR Egger method, which allows for an additional intercept (alpha) term which also provides an estimate of directional horizontal pleiotropy. This method relies upon the assumption that the size of the direct effects of the genetic variants on the outcome that do not operate through the exposure are independent of the variant’s effect on the exposure. Given possible instability in MR Egger estimates [35], we also used the bootstrap MR Egger method to meta-analyze the causal effect estimates from each SNP instrument. Further, we used four additional meta-analysis methods known to be more robust to presence of horizontal pleiotropy (at the expense of statistical power): penalised weighted median, simple mode, weighted median, and weighted mode [36].

Second, we restricted our choices of SNPs to those whose closest gene is directly involved in the vitamin D pathway. These genes have an established role in vitamin D regulation through its synthesis (*DHCR7*/*NADSYN1* and *CYP2R1*), transportation (*GC*), and degradation (*CYP24A1*) (**Supplement 3**). This decreases the risk of selecting a genetic variant that affects COVID-19 outcomes independent of its effect on 25OHD levels.

Third, we used the Phenoscanner tool [37,38] on the remaining SNPs to check for variants associated (at a genome-wide significant threshold of p=5×10^−8^) with phenotypes at risk of affecting COVID-19 outcomes independent of 25OHD, making them at higher risk of horizontal or vertical pleiotropy. Note that vertical pleiotropy, which happens when the COVID-19 outcome is influenced by a phenotype directly in the causal pathway between 25OHD level and COVID-19 outcome, does not violate MR assumptions.

### Research Ethics

Each cohort included in this study received their respective institutional research ethics board approval to enroll patients. All information used for this study are publicly available as deidentified GWAS summary statistics.

## Results

### Choice of 25OHD genetic instruments

We obtained our 25OHD genetic instruments from our previously published GWAS on circulating 25OHD levels in 401,460 white British participants in the UK Biobank (UKB) [39], which was meta-analyzed with a GWAS on 25OHD levels of 42,274 participants of European ancestry [40]. Of the 138 reported conditionally independent SNPs (explaining 4.9% of the 25OHD variance), 100 had a minor allele frequency of more than 1%, of which 78 were directly available in the COVID-19 HGI GWAS summary statistic and had linkage disequilibrium coefficient of less than 5%. Additionally, 3 more variants had good genetic proxies (r^2^>90%) and were therefore added to our instrument lists, for a total of 81 variants. These explained 4.3% of the variance in 25OHD serum levels. The full list of SNPs used can be found in **Supplement 4**.

### COVID-19 outcome definitions and GWASs

Using the COVID-19 HGI results restricted to cohorts of European ancestry, we used a total of 14,134 cases and 1,284,876 controls to define COVID-19 susceptibility, 6,406 cases and 902,088 controls to define COVID-19 hospitalization, and 4,336 cases and 623,902 controls to define COVID-19 severe disease. **Table 1** summarizes the definition and sample size of both the exposure and outcome GWASs. Since the UKB was used in the two phases of the MR study, some overlap between the exposure and the outcome GWASs was unavoidable (**Supplement 2**).

### Primary MR analysis

We first used IVW meta-analysis to combine effect estimates from each genetic instrument. For a standard deviation increase in log-transformed 25OHD level, we observed no statistically significant effect upon odds of susceptibility (OR = 0.97; 95% CI: 0.85, 1.10; P = 0.61). Of note, in the UKB, the distribution of 25OHD levels has a mean of 48.6 nmol/L and a standard deviation of 21.1 nmol/L. This standard deviation is comparable to what can be achieved with vitamin D supplementation, especially over short therapeutic courses [41]. Similarly, we observed no significant difference in risk of hospitalization (OR = 1.11; 95% CI: 0.91, 1.35; P = 0.30) or risk of severe disease (OR = 0.93; 95% CI: 0.73, 1.17; P = 0.53) associated with a standard deviation increase in log-transformed 25OHD level (**Table 2** and **Figure 1**).

**Table 2:** MR results. SNP: single nucleotide polymorphism. nSNPs: number of SNPs retained for this analysis. IVW: inverse-variance weighted method. CI: confidence interval. Confidence intervals were obtained using Normal approximations, explaining minor discrepancies with p-values close to the alpha=5% statistical significance threshold.

| Outcome | nSNPs | IVW OR (95% CI) | IVW p-value | IVW SNP Heterogeneity p-value | Egger alpha | Alpha p-value |
| --- | --- | --- | --- | --- | --- | --- |
| **25OHD primary analysis with all SNPs** |  |  |  |  |  |  |
| Susceptibility | 81 | 0.97 (0.85, 1.10) | 0.61 | 0.008 | 0.003 (-0.004, 0.009) | 0.43 |
| Hospitalization | 81 | 1.11 (0.91, 1.35) | 0.30 | 0.066 | 0.0002 (-0.010, 0.011) | 0.97 |
| Severe disease | 81 | 0.93 (0.73, 1.17) | 0.53 | 0.126 | 0.010 (-0.002, 0.022) | 0.11 |
| **25OHD sensitivity analysis restricted to genes in the vitamin D pathway** |  |  |  |  |  |  |
| Susceptibility | 12 | 0.96 (0.83, 1.11) | 0.59 | 0.160 | 0.005 (-0.021, 0.032) | 0.70 |
| Hospitalization | 12 | 1.07 (0.78, 1.47) | 0.67 | 0.004 | 0.031 (-0.027, 0.088) | 0.32 |
| Severe disease | 12 | 0.87 (0.64, 1.19) | 0.38 | 0.105 | 0.056 (0.006, 0.106) | 0.05 |
| **25OHD sensitivity analysis after removal of SNPs identified by Phenoscanner** |  |  |  |  |  |  |
| Susceptibility | 10 | 0.97 (0.77, 1.23) | 0.80 | 0.083 | 0.004 (-0.032, 0.040) | 0.83 |
| Hospitalization | 10 | 1.09 (0.68, 1.75) | 0.71 | 0.010 | 0.014 (-0.059, 0.088) | 0.71 |
| Severe disease | 10 | 0.91 (0.54, 1.55) | 0.73 | 0.095 | 0.072 (-0.003, 0.146) | 0.01 |

**Figure 1:**
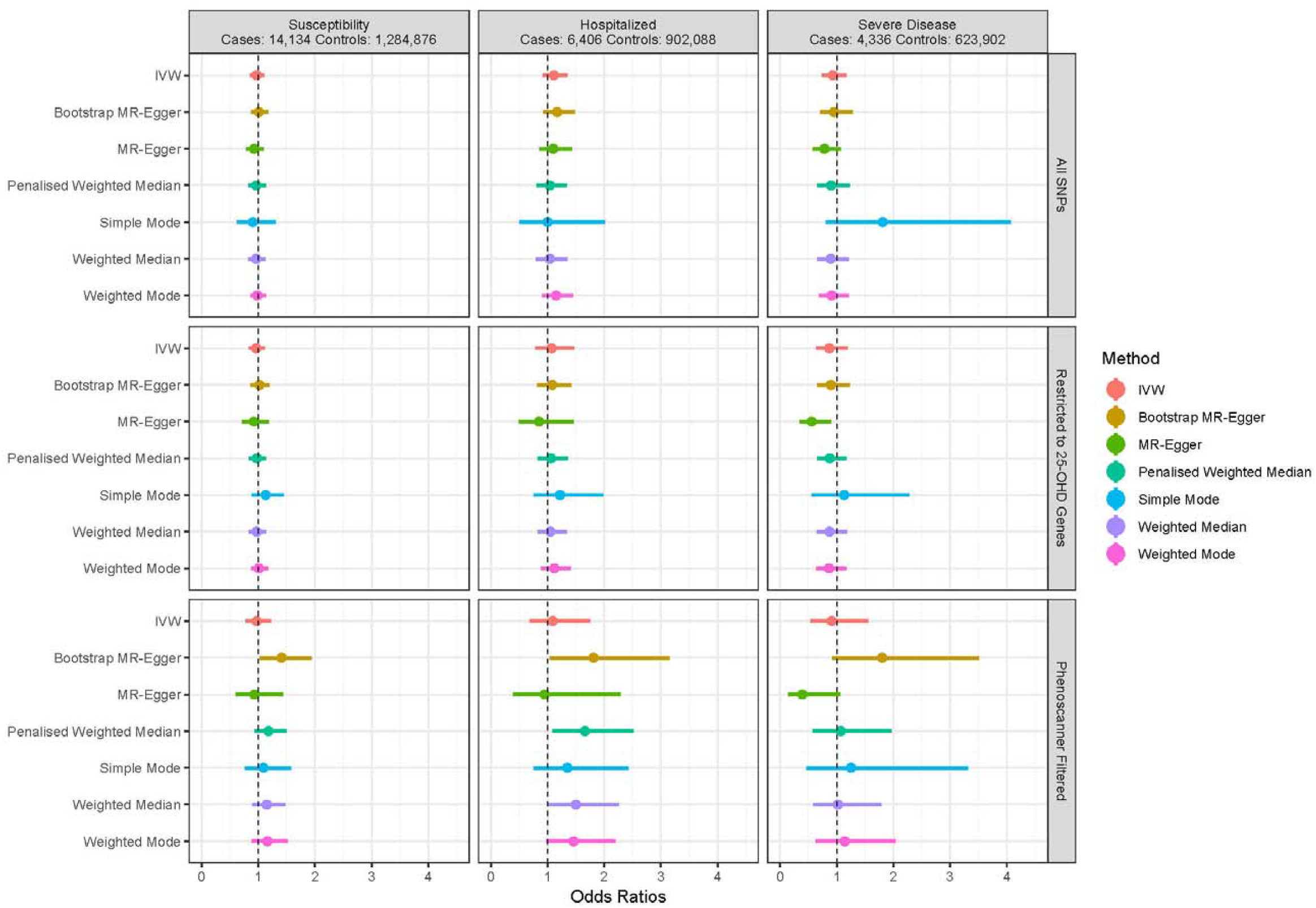
Odds ratio point estimates and 95% confidence intervals for a one standard deviation increase in 25OHD levels (on the log scale) on COVID-19 susceptibility and severity. Restricted to 25-OHD Genes: analysis restricted to SNPs near the 4 genes involved in known vitamin D metabolic pathways. Phenoscaner Filtered: analysis restricted to the 4 genes above, and with removal of SNPs identified to have other associations in Phenoscanner. Full results including odds ratios, confidence intervals, and p-values are available in **Supplement 5**.

### Horizontal pleiotropy assessment and sensitivity analysis

Using the MR Egger intercept terms, we did not observe evidence of horizontal pleiotropy. While they have less statistical power than IVW meta-analysis, the 6 sensitivity meta-analyses we used also showed no evidence of an association between 25OHD levels and COVID-19 susceptibility, hospitalization, and severe disease, with each confidence interval crossing the null in the primary analysis using all SNPs (**Figure 1** and **Supplement 5**). Our results are therefore unlikely to be strongly biased by horizontal pleiotropy.

Second, we restricted our analysis to SNPs which reside close to the four genes directly involved in 25OHD metabolism. This left 12 SNPs, explaining 3.2% of 25OHD variation. Using IVW, each standard deviation increase in log-transformed 25OHD was again not associated with COVID-19 susceptibility (OR = 0.96; 95% CI: 0.83, 1.11; P = 0.59), hospitalization (OR = 1.07 [95% CI: 0.78, 1.47]; P = 0.67) and severe disease (OR = 0.87; 95% CI: 0.63, 1.19; P = 0.38). For the three phenotypes, the MR Egger intercept term did not support bias from directional horizontal pleiotropy.

Lastly, we used the Phenoscanner [37,38] tool to check if the SNPs used in the MR study were associated with other phenotypes. Using Phenoscanner, rs11723621 was associated with white blood cell level, and rs6127099 was associated with glomerular filtration rate [42,43]. In both cases, the association with each phenotype was mild compared to their effect on 25OHD level, as rs11723621 explained less than 0.03% of the variance in white blood cell counts, and rs6127099 explained less than 0.001% of the glomerular filtration rate variance. Removing these SNPs from the 12 SNPs above further decreased the proportion of 25OHD variance explained to 1.7%. While confidence intervals widened, effect estimates when restricting our analysis to these SNPs remained null for susceptibility (0.80; 95% CI: 0.77, 1.23; P=0.80), hospitalization (1.09; 95% CI: 0.68, 1.75; P=0.71), and severe disease (0.91; 95% CI: 0.54, 1.55; P=0.73).

### Genetic instruments heterogeneity

Overall, our results showed little evidence of heterogeneity of effect between our genetic instruments (**Table 2**). We nonetheless observed that for at least one of the three analyses, we would have rejected the null hypothesis of homogeneous genetic effects in the COVID-19 hospitalization phenotype. However, given the large number of hypotheses tested, this may be due to chance.

## Discussion

In this large-scale MR study, we did not find evidence to support increasing 25OHD levels in order to protect against COVID-19 susceptibility, hospitalization, or severity. This lack of evidence was consistent across phenotypes, sensitivity analyses, and choice of genetic instruments. Differences between our findings and those reported in observational studies [6] may reflect the fact that associations between vitamin D and COVID-19 may be confounded due to factors difficult to control for even with advanced statistical adjustments, such as socio-economic status, institutionalizaton or medical comorbidities associated with lower vitamin D levels. While our study assessed the association between genetically determined levels of 25OHD and COVID-19, these results can still inform us on the role of vitamin D supplementation. Specifically, in contrast to observational studies, our findings do not support an association between higher 25OHD level and better COVID-19 outcome, and therefore do not support the use of vitamin D supplementation to prevent COVID-19 outcomes. Further, while a randomized trial [15] showed benefit of vitamin D supplementation using an endpoint at risk of bias due to the unblinded intervention (admission to the critical care unit) and a small sample size (n=75), a larger randomized trial [16] of 240 patients showed no effect of a single high dose of vitamin D3 on mortality, length of stay, or risk of mechanical ventilation. Thus, findings from the largest randomized trial to date are thus concordant with our MR results.

Our study’s main strength is MR’s track record of predicting RCT outcomes for multiple medical medical conditions [9–11,21–24,44,45]. Our study also leverages the largest cohort of COVID-19 cases and controls currently available (even outside of genetic studies) and the largest study on genetic determinants of 25OHD levels to date. Using these data sources, we were able to obtain results robust to multiple sensitivity analysis.

Our study still has limitations. First, our results do not apply to individuals with vitamin D deficiency, and it remains possible that truly deficient patients may benefit from supplementation for COVID-19 related protection and outcomes. However, individuals who are found to have frank vitamin D deficiency, should undergo replacement for bone protection. Second, our study may suffer from weak instrument bias, especially within sensitivity analyses that restricted to smaller sets of genetic instruments. In two-sample MR, this bias would tend to make estimates closer to the null. Nonetheless, similar studies have been able to use MR to establish an association between 25OHD levels and other diseases (most notably multiple sclerosis [25]), suggesting that these instruments are strong enough to find such associations. Further, given the large percentage of shared individuals from the UKB between the vitamin D exposure GWAS [28] and the severe COVID-19 phenotype, this analysis is close to a one-sample MR, which would show bias towards the observational study association. Given that this analysis also shows largely null effects, we do not suspect that weak instruments bias is a significant issue in our results. Third, given that vitamin D levels are affected by season (with higher levels after sunlight exposure), even if our SNP-instruments were obtained from a GWAS that controlled for season of blood draw, effect attenuation by averaging the effect of 25OHD levels on COVID-19 over all seasons may influence results. Nevertheless, a recent study in a Finnish cohort (where sun exposure greatly varies by season) showed that genetic determinants of 25OHD level were able to discriminate between individuals with predisposition to varying levels of 25OHD, regardless of the season [46]. Therefore, while the cyclical nature of 25OHD level is not completely modelled by MR, the size of this bias is likely small. Fourth, our MR analyses assume a linear exposure-outcome relationship. While this may slightly bias our results, simulation studies have previously shown that this assumption provides adequate results when looking at a population effect [27]. Therefore, for the purpose of vitamin D supplementation in the general population, our conclusions should still be valid. However, as pointed out above, we are not able to test the effect of vitamin D deficiency on COVID-19 outcomes. Lastly, as we only studied the effect of 25OHD and COVID-19 in individuals of European ancestry, it remains possible that 25OHD levels might have different effects on COVID-19 outcomes in other populations. However, previous RCTs on vitamin D supplementation have given similar results in populations of various ancestries [44,45].

In conclusion, using a method that has consistently replicated RCT results from vitamin D supplementation studies in large sample sizes, we find no evidence to support a protective role for higher 25OHD on COVID-19 outcomes. Specifically, vitamin D supplementation as a public health measure to improve COVID-19 outcomes is not supported by this MR study. Most importantly, our results suggest that investment in other therapeutic or preventative avenues should be prioritized for COVID-19 RCTs.

## Data Availability

Covid-19 outcome GWAS summary statistics are freely available for download directly through the Covid-19 HGI website (https://www.covid19hg.org/results/). The October 20th data freeze (v4) summary statistics were used for our study.

https://www.covid19hg.org/

## Acknowledgements

We thank the patients and investigators who contributed to the COVID-19 HGI (**Supplement 6**) and the Vitamin D GWAS consortium. Members of the GEN-COVID study are acknowledged in **Supplement 7**. This research has been conducted using the UK Biobank Resource (project number: 27449).

## Contributions

Conception and design: GBL, TN, JBR. Data acquisition and standardization: AR, AG, DRM, TA, OA, NM, NK, ZA. Data analyses: GBL and TN. Interpretation: GBL, TN, VM, DRM, TA, OA, NM, NK, ZA, AR, AG, SZ, YC, VF, JBR. Computational resources and support: VF, JBR. Writing original draft: GBL, TN, JBR. All authors were involved in reviewing the manuscript and critically reviewed its content. All authors gave final approval of the version to be published. The corresponding author attests that all listed authors meet authorship criteria and that no others meeting the criteria have been omitted.

## Supplementary files captions

**Supplement 1**: STROBE case-control study checklist

**Supplement 2**: Cohorts used for each outcome phenotype for the COVID-19 Host Genetics Initiative.

**Supplement 3**: Vitamin D metabolism pathway and genes involved.

**Supplement 4**: Genetic instruments summary statistics.

**Supplement 5**: results from Mendelian randomization sensitivity analyses.

**Supplement 6**: Acknowledgement to data contributors and the COVID-19 Host Genetics Initiative.

**Supplement 7**: GEN-COVID Multicenter Study.

