## Supplement 1 for "Vitamin D and COVID-19 susceptibility and severity in the COVID-19 Host Genetics Initiative: A Mendelian randomization study"

STROBE Statement—Checklist of items that should be included in reports of ***case-control studies***

|  | Item No | Recommendation |
| --- | --- | --- |
| **Title and abstract** | 1 | (*a*) Indicate the study’s design with a commonly used term in the title or the abstract  *Mendelian randomization is specified in the title and in the abstract methods.* |
|  |  | (*b*) Provide in the abstract an informative and balanced summary of what was done and what was found  *Provided in the abstract.* |
| Introduction | | |
| Background/rationale | 2 | Explain the scientific background and rationale for the investigation being reported  *Explained in paragraphs 1-5 of the introduction.* |
| Objectives | 3 | State specific objectives, including any prespecified hypotheses  *Stated in paragraph 6 of introduction.* |
| Methods | | |
| Study design | 4 | Present key elements of study design early in the paper  *Presented in the first paragraph of the methods.* |
| Setting | 5 | Describe the setting, locations, and relevant dates, including periods of recruitment, exposure, follow-up, and data collection  *Available information on vitamin D GWAS is provided in the “Choice of 25OHD genetic instruments” section.*  *Available information from the Covid-19 Host Genetics Initiative is provided in the “Covid-19 outcome definitions and GWASs” section of the methods.*  *Table 1 and Supplements 2 and 4 also provide more details on the exposures and outcomes.* |
| Participants | 6 | (*a*) Give the eligibility criteria, and the sources and methods of case ascertainment and control selection. Give the rationale for the choice of cases and controls  *This is described in detail in table 1 and in the “Covid-19 outcome definitions and GWASs” section.* |
|  |  | (*b*) For matched studies, give matching criteria and the number of controls per case  *Does not apply.* |
| Variables | 7 | Clearly define all outcomes, exposures, predictors, potential confounders, and effect modifiers. Give diagnostic criteria, if applicable  *Genetic instruments for vitamin D are described in the “Choice of 25OHD genetic instruments” and Supplement 4.*  *Outcomes are described in the “Covid-19 outcome definitions and GWASs” section and in table 1.* |
| Data sources/ measurement | 8* | For each variable of interest, give sources of data and details of methods of assessment (measurement). Describe comparability of assessment methods if there is more than one group  *Sources of GWASs are provided in the “Choice of 25OHD genetic instruments” and the “Covid-19 outcome definitions and GWASs” section, with reference or internet address.* |
| Bias | 9 | Describe any efforts to address potential sources of bias  *Horizonal pleiotropy assessment methods are described in the “Horizontal pleiotropy sensitivity analysis” section of the methods.* |
| Study size | 10 | Explain how the study size was arrived at  *This does not directly apply to a Mendelian randomization study, but the choice of genetic instruments was explained in the “Choice of 25OHD genetic instruments” section and in Supplement 4.* |
| Quantitative variables | 11 | Explain how quantitative variables were handled in the analyses. If applicable, describe which groupings were chosen and why  *Does not apply.* |
| Statistical methods | 12 | (*a*) Describe all statistical methods, including those used to control for confounding  *Mendelian randomization and sensitivity analyses are described in the “Primary MR analysis” and the “Horizontal pleiotropy sensitivity analysis” sections of the methods*. |
|  |  | (*b*) Describe any methods used to examine subgroups and interactions  *Does not apply*. |
|  |  | (*c*) Explain how missing data were addressed  *Does not apply.* |
|  |  | (*d*) If applicable, explain how matching of cases and controls was addressed  *Does not apply.* |
|  |  | (*e*) Describe any sensitivity analyses  *This was done in the “Horizontal pleiotropy sensitivity analysis” section.* |
| Results | | |
| Participants | 13* | (a) Report numbers of individuals at each stage of study—eg numbers potentially eligible, examined for eligibility, confirmed eligible, included in the study, completing follow-up, and analysed  *The number of individuals used for exposures and outcomes were given in the “Choice of 25OHD genetic instruments” and the “Covid-19 outcome definitions and GWASs” sections.*  *The number of genetic instruments was explained in the “Choice of 25OHD genetic instruments” and the “Horizontal pleiotropy assessment and sensitivity analysis” sections.* |
|  |  | (b) Give reasons for non-participation at each stage  *Does not directly apply to Mendelian randomization studies, but the reasons for excluding genetic variants was provided in the “Choice of 25OHD genetic instruments” and the “Horizontal pleiotropy assessment and sensitivity analysis” sections.* |
|  |  | (c) Consider use of a flow diagram  *Does not apply.* |
| Descriptive data | 14* | (a) Give characteristics of study participants (eg demographic, clinical, social) and information on exposures and potential confounders  *Available information was given in table 1, supplement 2, and the “Covid-19 outcome definitions and GWASs” section.* |
|  |  | (b) Indicate number of participants with missing data for each variable of interest  *Does not apply.* |
| Outcome data | 15* | Report numbers in each exposure category, or summary measures of exposure  *This was done in the “Covid-19 outcome definitions and GWASs” section, table 1, and supplement 2.* |
| Main results | 16 | (*a*) Give unadjusted estimates and, if applicable, confounder-adjusted estimates and their precision (eg, 95% confidence interval). Make clear which confounders were adjusted for and why they were included  *Confounder does not apply to Mendelian randomization, but primary MR analysis and horizontal pleiotropy analysis results are given in the “Primary MR analysis” and “Horizontal pleiotropy assessment and sensitivity analysis” sections of the results, as well as in figure 1.* |
|  |  | (*b*) Report category boundaries when continuous variables were categorized  *Does not apply.* |
|  |  | (*c*) If relevant, consider translating estimates of relative risk into absolute risk for a meaningful time period  *Does not apply.* |

| Other analyses | 17 | Report other analyses done—eg analyses of subgroups and interactions, and sensitivity analyses  *This was done in the “Horizontal pleiotropy assessment and sensitivity analysis” section of the result.* |
| --- | --- | --- |
| Discussion | | |
| Key results | 18 | Summarise key results with reference to study objectives  *This was done in the first paragraph of the discussion.* |
| Limitations | 19 | Discuss limitations of the study, taking into account sources of potential bias or imprecision. Discuss both direction and magnitude of any potential bias  *This was done in the third paragraph of the discussion.* |
| Interpretation | 20 | Give a cautious overall interpretation of results considering objectives, limitations, multiplicity of analyses, results from similar studies, and other relevant evidence  *This was done in the first paragraph of the discussion.* |
| Generalisability | 21 | Discuss the generalisability (external validity) of the study results  *This was done in paragraphs 1, 2, and 3 of the discussion.* |
| Other information | | |
| Funding | 22 | Give the source of funding and the role of the funders for the present study and, if applicable, for the original study on which the present article is based  *This is given in the “funding source” section.* |

*Give information separately for cases and controls.

**Note:** An Explanation and Elaboration article discusses each checklist item and gives methodological background and published examples of transparent reporting. The STROBE checklist is best used in conjunction with this article (freely available on the Web sites of PLoS Medicine at http://www.plosmedicine.org/, Annals of Internal Medicine at http://www.annals.org/, and Epidemiology at http://www.epidem.com/). Information on the STROBE Initiative is available at http://www.strobe-statement.org.
