## Supplement 2 for "Vitamin D and COVID-19 susceptibility and severity in the COVID-19 Host Genetics Initiative: A Mendelian randomization study"

**Supplement 1:** cohorts used for each outcome phenotype for the COVID-19 Host Genetics Initiative.

Each table is adapted from the COVID-19 Host Genetics Initiative website (Freeze 4: October 20, 2020). Available at: <https://www.covid19hg.org/>

Each cohort’s names are abbreviated in Tables 1.1-1.3. Full names are provided in Table 1.4 at the end of this supplement.

| **Cohort** | **Country** | **Ancestry** | **Cases** | **Controls** |
| --- | --- | --- | --- | --- |
| UKBB | UK | EUR | 1,305 | 370,602 |
| DECODE | Iceland | EUR | 1,897 | 273,257 |
| EstBB | Estonia | EUR | 313 | 138,272 |
| GENCOVID | Italy | EUR | 734 | 2,472 |
| FinnGen | Finland | FIN | 357 | 238,354 |
| genomicsengland100kgp | UK | EUR | 218 | 62,302 |
| SPGRX | Spain | EUR | 362 | 302 |
| Helix | USA | EUR | 178 | 5,441 |
| Lifelines | Netherlands | EUR | 358 | 25,213 |
| MGI | USA | EUR | 122 | 51,458 |
| MVP | USA | EUR | 1,520 | 7,600 |
| NTR | Netherlands | EUR | 145 | 5,252 |
| PHBB | USA | EUR | 151 | 29,966 |
| Stanford | USA | EUR | 109 | 191 |
| INTERVAL | UK | EUR | 161 | 41,674 |
| BQC19 | Canada | EUR | 206 | 327 |
| Ancestry | USA | EUR | 2,417 | 14,933 |
| Amsterdam_UMC_COVID_study_group | Netherlands | EUR | 108 | 1,413 |
| BelCovid | Belgium | EUR | 109 | 1,484 |
| HOSTAGE | Italy-Spain | EUR | 1,610 | 2,205 |
| SweCovid | Sweden | EUR | 78 | 3,778 |
| genomicc | UK | EUR | 1,676 | 8,380 |
| **Total** |  |  | 14,134 | 1,284,876 |

Table 1.1: Susceptibility phenotype

| **Cohort** | **Country** | **Ancestry** | **Cases** | **Controls** |
| --- | --- | --- | --- | --- |
| Amsterdam_UMC_COVID_study_group | Netherlands | EUR | 108 | 1,413 |
| DECODE | Iceland | EUR | 89 | 274,322 |
| BelCovid | Belgium | EUR | 109 | 1,484 |
| GENCOVID | Italy | EUR | 571 | 2,472 |
| FinnGen | Finland | FIN | 83 | 238,628 |
| SPGRX | Spain | EUR | 311 | 302 |
| HOSTAGE | Italy-Spain | EUR | 1,610 | 2,205 |
| BQC19 | Canada | EUR | 181 | 354 |
| UKBB | UK | EUR | 765 | 364,341 |
| MVP | USA | EUR | 436 | 2,180 |
| BoSCO | Germany | EUR | 139 | 262 |
| Ancestry | USA | EUR | 250 | 1,967 |
| SweCovid | Sweden | EUR | 78 | 3,778 |
| genomicc | UK | EUR | 1,676 | 8,380 |
| **Total** |  |  | 6406 | 902088 |

Table 1.2: Hospitalized phenotype

| **Cohort** | **Country** | **Ancestry** | **Cases** | **Controls** |
| --- | --- | --- | --- | --- |
| Amsterdam_UMC_COVID_study_group | Netherlands | EUR | 66 | 1,413 |
| BRACOVID | Brazil | AMR | 450 | 1,637 |
| GENCOVID | Italy | EUR | 468 | 2,472 |
| SweCovid | Sweden | EUR | 78 | 3,778 |
| FinnGen | Finland | FIN | 54 | 238,657 |
| genomicc | UK | EUR | 1,676 | 8,380 |
| SPGRX | Spain | EUR | 101 | 302 |
| BQC19 | Canada | EUR | 55 | 480 |
| UKBB | UK | EUR | 329 | 364,341 |
| Italy_HOSTAGE | Italy | EUR | 698 | 1,255 |
| Spain_HOSTAGE | Spain | EUR | 302 | 925 |
| BoSCO | Germany | EUR | 59 | 262 |
| **Total** |  |  | 4,336 | 623,902 |

Table 1.3: Severe disease phenotype

| **Full Name** | **Abbreviation** |
| --- | --- |
| Amsterdam UMC COVID study group | Amsterdam_UMC_COVID_study_group |
| AncestryDNA COVID-19 Research Study | Ancestry |
| Genetic determinants of COVID-19 complications in the Brazilian population | BRACOVID |
| Genetic modifiers for COVID-19 related illness | BelCovid |
| Biobanque Quebec COVID19 | BQC19 |
| deCODE | DECODE |
| Estonian Biobank | EstBB |
| FinnGen | FinnGen |
| GEN-COVID, reCOVID | GENCOVID |
| genomiCC | genomicc |
| COVID19-Host(a)ge | HOSTAGE |
| Helix Exome+ COVID-19 Phenotypes | Helix |
| UK Blood Donors Cohort | INTERVAL |
| Michigan Genomics Initiative | MGI |
| Million Veterans Program | MVP |
| Netherlands Twin Register | NTR |
| Partners Healthcare Biobank | PHBB |
| Determining the Molecular Pathways and Genetic Predisposition of the Acute Inflammatory Process Caused by SARS-CoV-2 | SPGRX |
| Genomic epidemiology of SARS-Cov-2 and host genetics in Coronavirus Disease 2019 (COVID-19) | Stanford |
| The genetic predisposition to severe COVID-19 | SweCovid |
| UK Biobank | UKBB |
| Bonn Study of COVID19 genetics | BoSCO |
| Italy COVID19-Host(a)ge | Italy_HOSTAGE |
| Spain COVID19-Host(a)ge | Spain_HOSTAGE |
| Genomics England | genomicsengland100kgp |
| Lifelines | Lifelines |

Table 1.4 Cohort name abbreviations
