## Supplement 3 for "Vitamin D and COVID-19 susceptibility and severity in the COVID-19 Host Genetics Initiative: A Mendelian randomization study"

**Supplement 2:** vitamin D synthesis pathway. For our sensitivity analyses, we restricted our instruments to variants in the circled genes below.

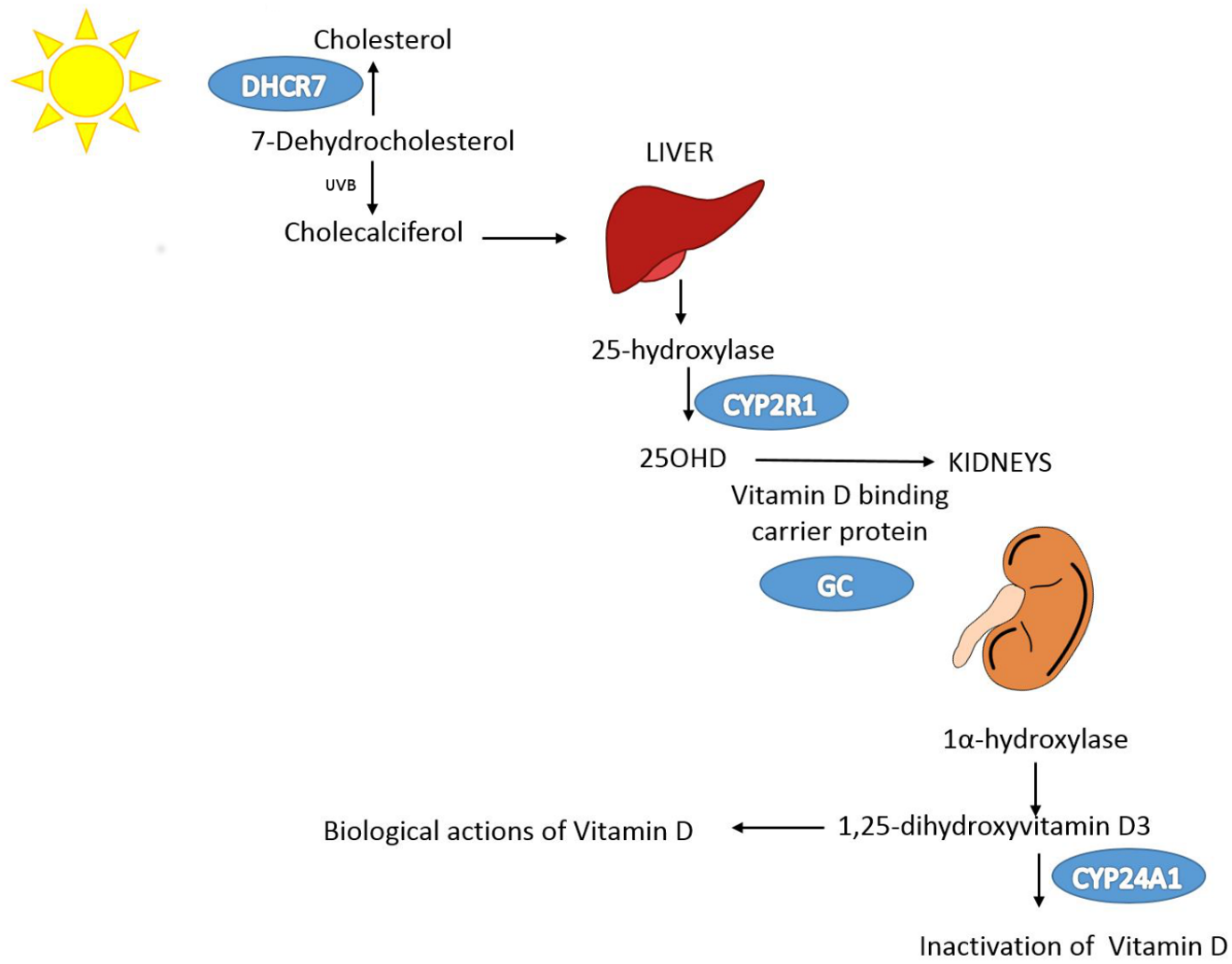
