## Supplement 4 for "Vitamin D and COVID-19 susceptibility and severity in the COVID-19 Host Genetics Initiative: A Mendelian randomization study"

**Supplement 3:** single nucleotide polymorphisms used as genetic instruments in each analysis

Genes in the vitamin D pathway are in bold.

| **SNP** | **CHR** | **BP** | **Effect Allele** | **Other Allele** | **Effect Allele Frequency** | **Beta** | **Standard error** | **P-value** | **Gene** |
| --- | --- | --- | --- | --- | --- | --- | --- | --- | --- |
| rs6698680 | 1 | 2329661 | G | A | 0.464195 | -0.01193 | 0.001947 | 8.99E-10 | RER1 |
| rs3750296 | 1 | 17559656 | C | G | 0.341423 | -0.02082 | 0.002042 | 2.09E-24 | PADI1 |
| rs7519574 | 1 | 34726552 | A | G | 0.181694 | 0.016991 | 0.002536 | 2.09E-11 | RP4-657M3.2 |
| rs56044892 | 1 | 41830086 | T | C | 0.2107 | 0.015388 | 0.00244 | 2.85E-10 | FOXO6 |
| rs2934744 | 1 | 63048045 | A | C | 0.643546 | -0.02241 | 0.002119 | 3.96E-26 | DOCK7 |
| rs7528419 | 1 | 109817192 | G | A | 0.224958 | 0.019031 | 0.002321 | 2.41E-16 | CELSR2 |
| rs3768013 | 1 | 150815411 | A | G | 0.369605 | -0.01488 | 0.002011 | 1.37E-13 | ARNT |
| rs115045402 | 1 | 152029548 | A | G | 0.026334 | 0.10713 | 0.006843 | 3.05E-55 | FLG |
| rs144613541 | 1 | 152270875 | G | A | 0.290985 | 0.015413 | 0.002244 | 6.49E-12 | FLG |
| rs61816761 | 1 | 152285861 | A | G | 0.023096 | 0.125479 | 0.006905 | 8.57E-74 | FLG |
| rs11264360 | 1 | 155284586 | A | T | 0.24292 | 0.018008 | 0.002286 | 3.34E-15 | FDPS |
| rs867772 | 1 | 220972343 | G | A | 0.681808 | -0.01384 | 0.002091 | 3.64E-11 | MARC_1 |
| rs12997242 | 2 | 21381177 | A | G | 0.437687 | -0.01251 | 0.001972 | 2.23E-10 | TDRD15 |
| rs11127048 | 2 | 27752463 | A | G | 0.616569 | 0.018107 | 0.002038 | 6.41E-19 | GCKR |
| rs6724965 | 2 | 101440151 | G | A | 0.171608 | -0.01654 | 0.002573 | 1.29E-10 | NPAS2 |
| rs7569755 | 2 | 118648261 | A | G | 0.292374 | 0.013923 | 0.002142 | 8.03E-11 | HTR5BP |
| rs1047891 | 2 | 211540507 | A | C | 0.316449 | -0.01417 | 0.002088 | 1.16E-11 | CPS1 |
| rs2011425 | 2 | 234627608 | G | T | 0.07939 | -0.04636 | 0.00361 | 9.66E-38 | UGT1A4 |
| rs1972994 | 3 | 85631142 | T | A | 0.64702 | -0.01751 | 0.002036 | 7.99E-18 | CADM2 |
| rs6438900 | 3 | 125148287 | G | C | 0.260706 | 0.013584 | 0.002221 | 9.59E-10 | MRPL3 |
| rs6773343 | 3 | 141825598 | T | C | 0.720232 | 0.01268 | 0.002171 | 5.20E-09 | TFDP2 |
| rs78649910 | 4 | 3482213 | A | T | 0.110004 | -0.01833 | 0.003122 | 4.32E-09 | DOK7 |
| rs7699711 | 4 | 69947596 | T | G | 0.454848 | -0.02864 | 0.001949 | 6.97E-49 | UGT2B7 |
| rs11723621 | 4 | 72615362 | G | A | 0.291123 | -0.18693 | 0.002121 | 3.0E-1443 | **GC** |
| rs58073039 | 4 | 88287363 | G | A | 0.298368 | -0.01412 | 0.002109 | 2.16E-11 | HSD17B11 |
| rs28364331 | 4 | 100201295 | G | A | 0.018845 | 0.061386 | 0.007186 | 1.31E-17 | ADH1A |
| rs1229984 | 4 | 100239319 | C | T | 0.973099 | -0.04685 | 0.006481 | 4.85E-13 | ADH1A |
| rs7718395 | 5 | 118652574 | G | C | 0.319522 | 0.012632 | 0.002096 | 1.67E-09 | TNFAIP8 |
| rs3822868 | 6 | 131934986 | G | A | 0.835029 | 0.021918 | 0.002745 | 1.41E-15 | MED23 |
| rs111529171 | 7 | 21571932 | C | G | 0.216376 | -0.01549 | 0.002369 | 6.24E-11 | DNAH11 |
| rs1011468 | 7 | 104613791 | A | G | 0.475748 | -0.0138 | 0.001946 | 1.35E-12 | LINC01004 |
| rs1858889 | 7 | 107117447 | C | A | 0.50084 | 0.012836 | 0.001942 | 3.85E-11 | COG5 |
| rs804280 | 8 | 11612698 | A | C | 0.581959 | 0.013033 | 0.001978 | 4.43E-11 | GATA4 |
| rs34726834 | 8 | 25889606 | T | C | 0.253615 | 0.013824 | 0.002239 | 6.65E-10 | EBF2 |
| rs7828742 | 8 | 116960729 | G | A | 0.596853 | -0.02193 | 0.00199 | 3.06E-28 | LINC00536 |
| rs10818769 | 9 | 125719923 | G | C | 0.856966 | -0.01697 | 0.00287 | 3.35E-09 | DNAH11 |
| rs532436 (proxy: rs635634) | 9 | 136149830 | A | G | 0.184 | -0.01524 | 0.002515 | 2.17E-09 | ABO |
| rs10887718 | 10 | 82042624 | T | C | 0.527362 | -0.01248 | 0.001946 | 1.44E-10 | MAT1A |
| rs10832218 | 11 | 14181174 | C | T | 0.19774 | -0.03422 | 0.002912 | 7.09E-32 | **CYP2R1** |
| rs577185477 | 11 | 14612563 | C | T | 0.01469 | -0.37937 | 0.009583 | 4.3E-311 | **CYP2R1** |
| rs10832289 | 11 | 14669496 | T | A | 0.410173 | -0.06852 | 0.001965 | 4.20E-233 | **CYP2R1** |
| rs188480917 (proxy: rs576128895) | 11 | 14785870 | G | C | 0.010804 | -0.34329 | 0.009688 | 3.10E-248 | **CYP2R1** |
| rs117576073 | 11 | 14912573 | T | G | 0.012385 | -0.11457 | 0.008813 | 1.22E-38 | **CYP2R1** |
| rs523583 | 11 | 66070146 | C | A | 0.469219 | 0.012174 | 0.001963 | 5.58E-10 | TMEM151A |
| rs12803256 | 11 | 71132868 | G | A | 0.770616 | 0.100325 | 0.002325 | 1.3E-378 | **DHCR7** |
| rs200454003 (proxy: rs4245442) | 11 | 71228990 | T | C | 0.264587 | -0.0867 | 0.002536 | 3.68E-256 | **DHCR7** |
| rs10793129 | 11 | 75459865 | A | G | 0.090078 | 0.024492 | 0.003468 | 1.64E-12 | RP11-21L23.4 |
| rs1149605 | 11 | 76485216 | C | T | 0.171453 | 0.01928 | 0.002577 | 7.34E-14 | RP11-21L23.4 |
| rs964184 | 11 | 116648917 | C | G | 0.863547 | 0.03977 | 0.002858 | 5.11E-44 | ZPR1 |
| rs2847500 | 11 | 120114421 | A | G | 0.124391 | -0.02113 | 0.002949 | 7.79E-13 | ZPR1 |
| rs12317268 | 12 | 21352541 | G | A | 0.151589 | -0.01853 | 0.002717 | 9.15E-12 | SLCO1B1 |
| rs9668081 | 12 | 38602911 | T | C | 0.47058 | 0.0116 | 0.001988 | 5.38E-09 | FAM166AP9 |
| rs10859995 | 12 | 96375682 | C | T | 0.581322 | -0.0394 | 0.001971 | 7.03E-89 | HAL |
| rs8018720 | 14 | 39556185 | C | G | 0.820235 | -0.03195 | 0.002546 | 4.04E-36 | SEC23A |
| rs261291 | 15 | 58680178 | C | T | 0.35583 | -0.02241 | 0.002033 | 2.89E-28 | LIPC |
| rs1800588 | 15 | 58723675 | T | C | 0.21461 | -0.02977 | 0.002366 | 2.65E-36 | LIPC |
| rs17765311 | 15 | 63789952 | C | A | 0.344611 | -0.01515 | 0.002047 | 1.35E-13 | AC007950.2 |
| rs62007299 | 15 | 77711719 | A | G | 0.709377 | -0.01444 | 0.002145 | 1.69E-11 | PEAK1 |
| rs8063706 | 16 | 11909552 | T | A | 0.272828 | 0.012968 | 0.002198 | 3.64E-09 | BCAR4 |
| rs77924615 | 16 | 20392332 | A | G | 0.197773 | -0.01579 | 0.002464 | 1.46E-10 | PDILT |
| rs1800775 | 16 | 56995236 | A | C | 0.486317 | -0.01662 | 0.00195 | 1.56E-17 | CETP |
| rs2909218 | 17 | 66464546 | T | C | 0.792744 | 0.016894 | 0.002418 | 2.81E-12 | RP11-120M18.2 |
| rs8091117 | 18 | 28919794 | A | C | 0.065396 | -0.02407 | 0.003943 | 1.03E-09 | DSG1 |
| rs2037511 | 18 | 61366207 | A | G | 0.165391 | 0.016025 | 0.002618 | 9.29E-10 | SERPINB11 |
| rs57631352 | 19 | 4338173 | G | A | 0.297255 | -0.01287 | 0.002128 | 1.48E-09 | STAP2 |
| rs73015021 | 19 | 11192915 | G | A | 0.121031 | 0.023034 | 0.002983 | 1.15E-14 | LDLR |
| rs10500209 | 19 | 11979164 | C | T | 0.282129 | -0.01342 | 0.002169 | 6.18E-10 | LDLR |
| rs58542926 | 19 | 19379549 | T | C | 0.075847 | 0.032488 | 0.00367 | 8.57E-19 | TM6SF2 |
| rs3814995 | 19 | 36342212 | T | C | 0.31224 | -0.01473 | 0.002109 | 2.83E-12 | NPHS1 |
| rs1065853 | 19 | 45413233 | T | G | 0.082083 | 0.027397 | 0.00367 | 8.32E-14 | APOC1 |
| rs157595 | 19 | 45425460 | G | A | 0.614127 | -0.01558 | 0.00205 | 2.95E-14 | APOC1 |
| rs112285002 (proxy: rs296386) | 19 | 48374320 | T | C | 0.159529 | 0.060321 | 0.002701 | 1.77E-110 | SULT2A1 |
| rs10426 | 19 | 51517798 | A | G | 0.213371 | 0.025226 | 0.002382 | 3.31E-26 | KLK10 |
| rs8103262 | 19 | 53065814 | C | T | 0.305004 | 0.012519 | 0.002114 | 3.18E-09 | ZNF808 |
| rs6123359 | 20 | 52714706 | G | A | 0.105192 | 0.032345 | 0.003213 | 7.74E-24 | **CYP24A1** |
| rs6127099 | 20 | 52731402 | T | A | 0.279034 | -0.0368 | 0.002219 | 9.30E-62 | **CYP24A1** |
| rs2585442 | 20 | 52737123 | G | C | 0.246306 | 0.033609 | 0.002287 | 6.87E-49 | **CYP24A1** |
| rs2762942 | 20 | 52788925 | A | G | 0.942083 | 0.053192 | 0.004321 | 7.99E-35 | **CYP24A1** |
| rs2229742 | 21 | 16339172 | C | G | 0.103928 | -0.02574 | 0.00319 | 7.13E-16 | NRIP1 |
| rs2074735 | 22 | 31535872 | C | G | 0.064197 | 0.027256 | 0.003969 | 6.55E-12 | PLA2G3 |
| rs960596 | 22 | 41393520 | T | C | 0.339588 | 0.012414 | 0.002076 | 2.23E-09 | SCUBE1 |
