## Supplement 5 for "Vitamin D and COVID-19 susceptibility and severity in the COVID-19 Host Genetics Initiative: A Mendelian randomization study"

**Supplement 4**: Mendelian randomization sensitivity analyses results.

Odds ratios are given as the increase in odds of the Covid-19 outcome for every increase of one standard deviation log(25OHD). That is, an odds ratio greater than one indicates higher odds of the Covid-19 outcome. Confidence intervals are given using a Normal approximation, explaining minor discrepancies with p-values in cases close to the alpha=5% threshold of statistical significance.

| **Outcomes** | **Bootstrap MR Egger** | | **MR Egger** | | **Penalized Weighted Median** | | **Simple Mode** | | **Weighted Median** | | **Weighted Mode** | |
| --- | --- | --- | --- | --- | --- | --- | --- | --- | --- | --- | --- | --- |
|  | **OR (95% CI)** | **P** | **OR (95% CI)** | **P** | **OR (95% CI)** | **P** | **OR (95% CI)** | **P** | **OR (95% CI)** | **P** | **OR (95% CI)** | **P** |
| ***25OHD primary analysis with all SNPs*** | | | | | | | | | | | | |
| Susceptibility | 1.01 (0.872, 1.18) | 0.422 | 0.927 (0.786, 1.09) | 0.373 | 0.965 (0.823, 1.13) | 0.662 | 0.901 (0.61, 1.33) | 0.602 | 0.963 (0.823, 1.13) | 0.632 | 0.983 (0.861, 1.12) | 0.8 |
| Hospitalization | 1.16 (0.906, 1.49) | 0.122 | 1.1 (0.854, 1.43) | 0.449 | 1.04 (0.808, 1.34) | 0.754 | 1 (0.481, 2.09) | 0.994 | 1.04 (0.801, 1.35) | 0.778 | 1.15 (0.916, 1.44) | 0.235 |
| Severe Disease | 0.957 (0.72, 1.27) | 0.376 | 0.781 (0.572, 1.07) | 0.123 | 0.9 (0.653, 1.24) | 0.517 | 1.81 (0.787, 4.17) | 0.167 | 0.893 (0.671, 1.19) | 0.435 | 0.909 (0.688, 1.2) | 0.503 |
| ***25OHD sensitivity analysis restricted to genes in the vitamin D pathway*** | | | | | | | | | | | | |
| Susceptibility | 1.02 (0.866, 1.2) | 0.412 | 0.923 (0.716, 1.19) | 0.549 | 0.973 (0.835, 1.13) | 0.728 | 1.13 (0.882, 1.45) | 0.351 | 0.969 (0.829, 1.13) | 0.691 | 1.01 (0.873, 1.17) | 0.908 |
| Hospitalization | 1.08 (0.816, 1.42) | 0.289 | 0.849 (0.494, 1.46) | 0.566 | 1.06 (0.823, 1.36) | 0.661 | 1.22 (0.753, 1.98) | 0.437 | 1.05 (0.824, 1.34) | 0.696 | 1.12 (0.882, 1.41) | 0.381 |
| Severe Disease | 0.896 (0.655, 1.23) | 0.254 | 0.556 (0.343, 0.902) | 0.0386 | 0.876 (0.654, 1.17) | 0.374 | 1.13 (0.559, 2.28) | 0.742 | 0.872 (0.642, 1.18) | 0.38 | 0.863 (0.638, 1.17) | 0.36 |
| ***25OHD sensitivity analysis after removal of SNPs identified by Phenoscanner*** | | | | | | | | | | | | |
| Susceptibility | 1.41 (1.02, 1.94) | 0.017 | 0.932 (0.602, 1.44) | 0.761 | 1.18 (0.935, 1.5) | 0.161 | 1.09 (0.759, 1.58) | 0.641 | 1.15 (0.898, 1.48) | 0.263 | 1.16 (0.886, 1.52) | 0.308 |
| Hospitalization | 1.81 (1.04, 3.15) | 0.016 | 0.948 (0.392, 2.29) | 0.909 | 1.66 (1.09, 2.52) | 0.0171 | 1.35 (0.751, 2.43) | 0.342 | 1.5 (0.994, 2.26) | 0.0534 | 1.46 (0.981, 2.19) | 0.0949 |
| Severe Disease | 1.8 (0.917, 3.51) | 0.045 | 0.393 (0.146, 1.06) | 0.103 | 1.07 (0.581, 1.97) | 0.829 | 1.25 (0.469, 3.32) | 0.667 | 1.02 (0.584, 1.78) | 0.947 | 1.14 (0.631, 2.04) | 0.683 |
