## Supplement 6 for "Vitamin D and COVID-19 susceptibility and severity in the COVID-19 Host Genetics Initiative: A Mendelian randomization study"

**Supplement 5**: Acknowledgement to data contributors and the Covid-19 Host Genetics Initiative

**Covid-19 Host Genetics Initiative Coordination**

***Phenotype steering group***

Les Biesecker

Lea Davis

Patrick Deelen

Andrea Ganna

David van Heel

Eric Kerchberger

Sulggi Lee

Tomoko Nakanishi

James Priest

Alessandra Renieri

Brent Richards

Vijay Sankaran

***Administrative support***

Karolina Chwialkowska

Margherita Francescatto

Christine Stevens

***International Common Disease Alliance***

Amy Trankiem

Kate Balaconis

***Leadership***

Rachel Liao

Mark Daly

Andrea Ganna

Ben Neale

***Data dictionary***

Anna Bernasconi

Stefano Ceri

Francesca Mari

Alessandra Renieri

***Analysis***

Juha Karjalainen

Mattia Cordioli

Mari Niemi

Wei Zhou

***Website***

Huy Nguyen

Matthew Solomonson

**Data contributors, in cohort alphabetical order.**

**COVID19-Host(a)ge (Germany)**

***Ethics and communication***

Agustín Albillos

Rosanna Asselta

Luis Bujanda

Maria Buti

Stefano Duga

Javier Fernández

Manuel Romero Gomez

Pietro Invernizzi

Daniele Prati

***Data collection and coordination***

Jesus M. Banales

Trine Folseraas

Andre Franke

Johannes R Hov

Tom H Karlsen

Luca Valenti

***Analysis***

Frauke Degenhardt

David Ellighaus

**deCODE (Iceland)**

***Data collection and coordination***

Elias S Eythorsson

Asgeir Haraldsson

Dadi Helgason

Hilma Holm

Ragnar F Ingvarsson

Ingileif Jonsdottir

Gudmundur L Norddahl

Runolfur Palsson

Jona Saemundsdottir

Kari Stefansson

Unnur Thorsteinsdottir

***Analysis***

Daníel F Gudbjartsson

Hakon Jonsson

Pall Melsted

Patrick Sulem

Gardar Sveinbjornsson

**FinnGen (Finland)**

***Data collection and coordination***

Finngen

**Genetic modifiers for COVID-19 related illness (Belgium)**

***Data collection and coordination***

Adeline Busson

Jean-Christophe Goffard

Isabelle Migeotte

Xavier Peyrassol

Guillaume Smits

Isabelle Vandernoot

Francoise Wilkin

***Technical support***

Youssef Bouysran

Bruno Pichon

Nicky Tiembe

**GEN-COVID (Italy)**

See **Supplement 6**.

**Helix Exome+ (USA)**

***Data collection and coordination***

Kelly M. Schiabor Barrett

Alexandre Bolze

Elizabeth T. Cirulli

Jimmy M. Ramirez III

Yan Wei Lim

James T. Lu

Stephen Riffle

Francisco Tanudjaja

Xueqing Wang

Nicole L. Washington

Simon White

**LifeLines (Netherlands)**

***Analysis***

Annique Claringbould

Patrick Deelen

Esteban Lopera

Robert Warmerdam

***Data collection and coordination***

Marike Boezen

Lude Franke

**Netherlands Twin Register (Netherlands)**

***Data collection and coordination***

Meike Bartels

Eco de Geus

Michel G Nivard

***Analysis***

Jouke-Jan Hottenga

**Partners Healthcare Biobank, Greater Boston Covid-19 Host Disease Initiative, and Mass General Brigham – Host Vulnerability to COVID-19 (USA)**

***Data collection and coordination***

Robert Green

Beth Karlson

James Meigs

Josep Mercader

Shawn Murphy

Emma Perez

Sue Slaugenhaupt

Jordan Smoller

Scott Weiss

Ann Woolley

***Analysis***

Yen-Chen Anne Feng

***Other***

Ben Neal

Vijay G. Sankaran

**UK Biobank (UK)**

***Analysis***

Elizabeth G. Atkinson

Nikolas Baya

Guillaume Butler-Laporte

Hilary Finucane

Vincenzo Forgetta

Masahiro Kanai

Konrad J. Karczewski

Nils Koelling

Alicia R. Martin

Tomoko Nakanishi

Duncan S. Palmer

J. Brent Richards

Chris C A Spencer

Patrick Turley

Raymond K. Walters

Daniel J Wilson

***Data collection and coordination***

Jacob Armstrong

Anne Marie O'Connell

David H Wyllie

***Technical support***

Sam Bryant

***Administrative support***

Claire Churchhouse

**UK Blood Donors Cohort (UK)**

***Data collection and coordination***

Emanuele Di Angelantonio

Michael Chapman

John Danesh

Willem Ouwehand

Dave Roberts

Nick Watkins

***Analysis***

Adam Butterworth

Jing Hua Zhao

**UK 100,000 Genomes Project (UK)**

***Data collection and coordination***

Prabhu Arumugam

Mark Caulfield

Genomics England Research Consortium

Anna C Need

Thomas Oscroft

Augusto Rendon

Richard H Scott

***Analysis***

Georgia Chan

Athanasios Kousathanas

Loukas Moutsianas

Chris A Odhams

Dorota Pasko

Dan Rhodes

Alex Stuckey
