## Supplement 7 for "Vitamin D and COVID-19 susceptibility and severity in the COVID-19 Host Genetics Initiative: A Mendelian randomization study"

**Supplement 6:** GEN-COVID Multicenter Study **(**[**https://sites.google.com/dbm.unisi.it/gen-covid**](https://sites.google.com/dbm.unisi.it/gen-covid)**)**

Francesca Montagnani^3,5^, Chiara Fallerini^1^, Margherita Baldassarri^1^, Annarita Giliberti^1^, Elisa Benetti^3^, Floriana Valentino^1^, Sergio Daga^1^, Gabriella Doddato^1^, Susanna Croci^1^, Rossella Tita^2^, Francesca Fava^1,2^, Mirella Bruttini^1,2^, Elisa Frullanti^1^, Anna Maria Pinto^2^, Francesca Mari^1,2^, Simone Furini^3^, Laura Di Sarno^1^, Andrea Tommasi^1,2^, Maria Palmieri^1^, Arianna Emiliozzi^3,4^, Massimiliano Fabbiani^4^, Barbara Rossetti^4^, Giacomo Zanelli^3,4^, Laura Bergantini^5^, Miriana D’Alessandro^5^, Paolo Cameli^5^, David Bennet^5^, Federico Anedda^6^, Simona Marcantonio^6^, Sabino Scolletta^6^, Federico Franchi^6^, Maria Antonietta Mazzei^7^, Edoardo Conticini^8^, Luca Cantarini^8^, Bruno Frediani^8^, Danilo Tacconi^9^, Marco Feri^10^, Raffaele Scala^11^, Genni Spargi^12^, Marta Corridi^12^, Cesira Nencioni^13^, Gian Piero Caldarelli^14^, Maurizio Spagnesi^15^, Paolo Piacentini^15^ , Maria Bandini^15^, Elena Desanctis^15^, Anna Canaccini^16^, Chiara Spertilli^9^, Alice Donati^10^, Luca Guidelli^11^, Leonardo Croci^13^, Agnese Verzuri^16^, Valentina Anemoli^16^, Agostino Ognibene^17^, Massimo Vaghi^18^, Antonella D’Arminio Monforte^19^, Esther Merlini^19^, Mario U. Mondelli^20,21^, Stefania Mantovani^20^, Serena Ludovisi^20,21^, Massimo Girardis^22^, Sophie Venturelli^22^, Marco Sita^22^, Andrea Cossarizza^23^, Andrea Antinori^24^, Alessandra Vergori^24^, Stefano Rusconi^25,26^, Matteo Siano^26^, Arianna Gabrieli^26^, Agostino Riva^25,26^, Daniela Francisci^27,28^, Elisabetta Schiaroli^27^, Pier Giorgio Scotton^29^, Francesca Andretta^29^, Sandro Panese^30^, Renzo Scaggiante^31^, Saverio Giuseppe Parisi^32^, Francesco Castelli^33^, Maria Eugenia Quiros-Roldan^33^, Paola Magro^33^, Cristina Minardi^33^, Deborah Castelli^33^, Itala Polesini^33^, Matteo Della Monica^34^, Carmelo Piscopo^34^, Mario Capasso^35,36,37^, Roberta Russo^35,36^, Immacolata Andolfo^35,36^, Achille Iolascon^35,36^, Massimo Carella^38^, Marco Castori^38^, Giuseppe Merla^38^, Filippo Aucella^39^, Pamela Raggi^40^, Carmen Marciano^40^, Rita Perna^40^, Matteo Bassetti^41,42^, Antonio Di Biagio^42^, Maurizio Sanguinetti^43,44^, Luca Masucci^43,44^, Chiara Gabbi^45^, Serafina Valente^46^, Susanna Guerrini^8^, Ilaria Meloni^1^, Maria Antonietta Mencarelli^2^, Caterina Lo Rizzo^2^, Elena Bargagli^6^, Marco Mandalà^47^, Alessia Giorli^47^, Lorenzo Salerni^47^, Giuseppe Fiorentino^48^, Patrizia Zucchi^49^, Pierpaolo Parravicini^49^, Elisabetta Menatti^50^, Stefano Baratti^51^, Tullio Trotta^52^, Ferdinando Giannattasio^52^, Gabriella Coiro^52^, Fabio Lena^53^, Domenico A. Coviello^54^, Cristina Mussini^55^

1. Medical Genetics, University of Siena, Italy
2. Genetica Medica, Azienda Ospedaliera Universitaria Senese, Italy
3. Dept of Medical Biotechnologies, University of Siena, Italy
4. Dept of Specialized and Internal Medicine, Tropical and Infectious Diseases Unit
5. Unit of Respiratory Diseases and Lung Transplantation, Department of Internal and Specialist Medicine, University of Siena
6. Dept of Emergency and Urgency, Medicine, Surgery and Neurosciences, Unit of Intensive Care Medicine, Siena University Hospital, Italy
7. Department of Medical, Surgical and Neuro Sciences and Radiological Sciences, Unit of Diagnostic Imaging, University
8. Rheumatology Unit, Department of Medicine, Surgery and Neurosciences, University of Siena, Policlinico Le Scotte, Italy
9. Department of Specialized and Internal Medicine, Infectious Diseases Unit, San Donato Hospital Arezzo, Italy
9. Dept of Emergency, Anesthesia Unit, San Donato Hospital, Arezzo, Italy
9. Department of Specialized and Internal Medicine, Pneumology Unit and UTIP, San Donato Hospital, Arezzo, Italy
9. Department of Emergency, Anesthesia Unit, Misericordia Hospital, Grosseto, Italy
9. Department of Specialized and Internal Medicine, Infectious Diseases Unit, Misericordia Hospital, Grosseto, Italy
9. Clinical Chemical Analysis Laboratory, Misericordia Hospital, Grosseto, Italy
9. Department of Prevention, Azienda USL Toscana Sud Est, Italy
9. Territorial Scientific Technician Department, Azienda USL Toscana Sud Est, Italy
9. Clinical Chemical Analysis Laboratory, San Donato Hospital, Arezzo, Italy
18. Chirurgia Vascolare, Ospedale Maggiore di Crema, Italy
18. Department of Health Sciences, Clinic of Infectious Diseases, ASST Santi Paolo e Carlo, University of Milan, Italy
18. Division of Infectious Diseases and Immunology, Fondazione IRCCS Policlinico San Matteo, Pavia, Italy
18. Department of Internal Medicine and Therapeutics, University of Pavia, Italy
18. Department of Anesthesia and Intensive Care, University of Modena and Reggio Emilia, Modena, Italy
18. Department of Medical and Surgical Sciences for Children and Adults, University of Modena and Reggio Emilia,

Modena, Italy

18. HIV/AIDS Department, National Institute for Infectious Diseases, IRCCS, Lazzaro Spallanzani, Rome, Italy
18. III Infectious Diseases Unit, ASST-FBF-Sacco, Milan, Italy
18. Department of Biomedical and Clinical Sciences Luigi Sacco, University of Milan, Milan, Italy
18. Infectious Diseases Clinic, Department of Medicine 2, Azienda Ospedaliera di Perugia and University of Perugia,

Santa Maria Hospital, Perugia, Italy

18. Infectious Diseases Clinic, "Santa Maria" Hospital, University of Perugia, Perugia, Italy
18. Department of Infectious Diseases, Treviso Hospital, Local Health Unit 2 Marca Trevigiana, Treviso, Italy
18. Infectious Diseases Department, Ospedale Civile "SS. Giovanni e Paolo", Venice, Italy
18. Infectious Diseases Clinic, ULSS1, Belluno, Italy
18. Department of Molecular Medicine, University of Padova, Italy
18. Department of Infectious and Tropical Diseases, University of Brescia and ASST Spedali Civili Hospital, Brescia, Italy
18. Medical Genetics and Laboratory of Medical Genetics Unit, A.O.R.N. "Antonio Cardarelli", Naples, Italy
18. Department of Molecular Medicine and Medical Biotechnology, University of Naples Federico II, Naples, Italy
18. CEINGE Biotecnologie Avanzate, Naples, Italy
18. IRCCS SDN, Naples, Italy
18. Division of Medical Genetics, Fondazione IRCCS Casa Sollievo della Sofferenza Hospital, San Giovanni Rotondo,

Italy

18. Department of Medical Sciences, Fondazione IRCCS Casa Sollievo della Sofferenza Hospital, San Giovanni

Rotondo, Italy

18. Clinical Trial Office, Fondazione IRCCS Casa Sollievo della Sofferenza Hospital, San Giovanni Rotondo, Italy
18. Department of Health Sciences, University of Genova, Genova, Italy
18. Infectious Diseases Clinic, Policlinico San Martino Hospital, IRCCS for Cancer Research Genova, Italy
18. Microbiology, Fondazione Policlinico Universitario Agostino Gemelli IRCCS, Catholic University of Medicine,

Rome, Italy

18. Department of Laboratory Sciences and Infectious Diseases, Fondazione Policlinico Universitario A. Gemelli IRCCS,

Rome, Italy

18. Independent Scientist, Milan, Italy
18. Department of Cardiovascular Diseases, University of Siena, Siena, Italy
18. Otolaryngology Unit, University of Siena, Italy
18. AORN dei Colli Presidio Ospedaliero Cotugno, Italy
18. Department of Internal Medicine, ASST Valtellina e Alto Lario, Sondrio, Italy
18. Study Coordinator Oncologia Medica e Ufficio Flussi Sondrio, Italy
18. Department of Infectious and Tropical Diseases, University of Padova, Padova, Italy
18. First Aid Department, Luigi Curto Hospital, Polla, Salerno, Italy
18. Local Health Unit-Pharmaceutical Department of Grosseto, Toscana Sud Est Local Health Unit, Grosseto, Italy
18. U.O.C. Laboratorio di Genetica Umana, IRCCS Istituto G. Gaslini, Genoa, Italy.
18. Infectious Diseases Clinics, University of Modena and Reggio Emilia, Modena, Italy.
